# Phenotype Execution and Modelling Architecture (PhEMA) to support disease surveillance and real-world evidence studies: English sentinel network evaluation

**DOI:** 10.1101/2023.11.21.23298758

**Authors:** Gavin Jamie, William Elson, Simon de Lusignan, Debasish Kar, Rashmi Wimalaratna, Uy Hoang, Bernardo Meza-Torres, Anna Forbes, Will Hinton, Sneha Anand, Filipa Ferreira, Jose Ordonez-Mena, Utkarsh Agrawal, Rachel Byford

## Abstract

**Objective:** To evaluate Phenotype Execution and Modelling Architecture (PhEMA), to express sharable phenotypes using Clinical Query Language (CQL) and intensional SNOMED CT Fast Healthcare Interoperability Resources (FHIR) valuesets, for exemplar chronic disease, sociodemographic risk factor and surveillance phenotypes.

**Method:** We curated three phenotypes: Type 2 diabetes (T2DM), excessive alcohol use and incident influenza-like illness (ILI) using CQL to define clinical and administrative logic. We defined our phenotypes with valuesets, using SNOMED’s hierarchy and expression constraint language (ECL), and CQL, combining valuesets and adding temporal elements where needed. We compared the count of cases found using PhEMA with our existing approach using convenience datasets.

**Results:** The T2DM phenotype could be defined as two intensionally defined SNOMED valuesets and a CQL script. It increased the prevalence from 7.2% to 7.3%. Excess alcohol phenotype was defined by valuesets that added qualitative clinical terms to the quantitative conceptual definitions we currently use; this change increased prevalence by 58%, from 1.2% to 1.9%. We created an ILI valueset with SNOMED concepts, adding a temporal element using CQL to differentiate new episodes. This increased the weekly incidence in our convenience sample (weeks 26 to 38) from 0.95 cases to 1.11 cases per 100,000 people.

**Conclusions:** Phenotypes for surveillance and research can be described fully and comprehensibly using CQL and intensional FHIR valuesets. Our use case phenotypes identified a greater number of cases, whilst anticipated from excessive alcohol this was not for our other variable. This may have been due to our use of SNOMED CT hierarchy.

## Introduction

### Moving towards shareable phenotypes

A digital phenotype refers to the rules that are applied to computerised medical records (CMR) to identify cohorts of patients or episodes of interest.[1] The rules are converted to computer algorithms that typically include both logical expressions and data elements such as lists of clinical codes. These algorithms are applied through computerised queries to a computerised medical record (CMR) system or other health data repository to return the desired data output.[2]

Phenotypes should be transferable between organisations and environments, to support the goals of open science.[3] To ensure interpretability and facilitate electronic sharing of algorithms within data pipelines phenotypes should be represented in human and machine-readable formats. A number of public phenotype libraries have been developed to support the sharing and distribution of algorithms. Fourteen desiderata have been identified for the ideal phenotype library, though they are rarely even partially met.[4]

### Digital phenotyping at the Oxford-Royal College of General Practitioners (RCGP)-Research and Surveillance Centre (RSC)

The RSC collects CMR data from over 1,800 English primary care practices for surveillance, research, quality improvement, and educational purposes.[5] This pseudonymised data is securely stored in a trusted research environment (TRE).[6] The RSC has issued weekly disease surveillance reports to the UK Health Security Agency (UKHSA) and its predecessors for over 50 years, and conducts chronic disease CMR research. Development of digital phenotypes, or phenotyping, is pivotal for the work undertaken by the RSC.[7]

At the RSC, we use a three-layered approach to phenotyping shown in Figure 1.[8] Ontological layer: a human-readable description of the key concepts and relationships of a phenotype. Coding layer: where the conceptual reasoning is used to generate lists of clinical codes. Our code lists are typically based on Systematised Nomenclature of Medicine (SNOMED) Clinical Terms (CT) as this is the mandated terminology used in UK primary care.[9] SNOMED CT code lists are typically formatted as refsets. We develop refsets using our in house software tool, the “SNOMED helper tool”, shown in Supplementary Figure S1. Logical data extract layer: where Structured Query Language (SQL) queries are manually written using logical expressions combined with the refsets to extract the desired data. The data is subsequently tested for face validity and compared with other data sources.

**Figure 1:**
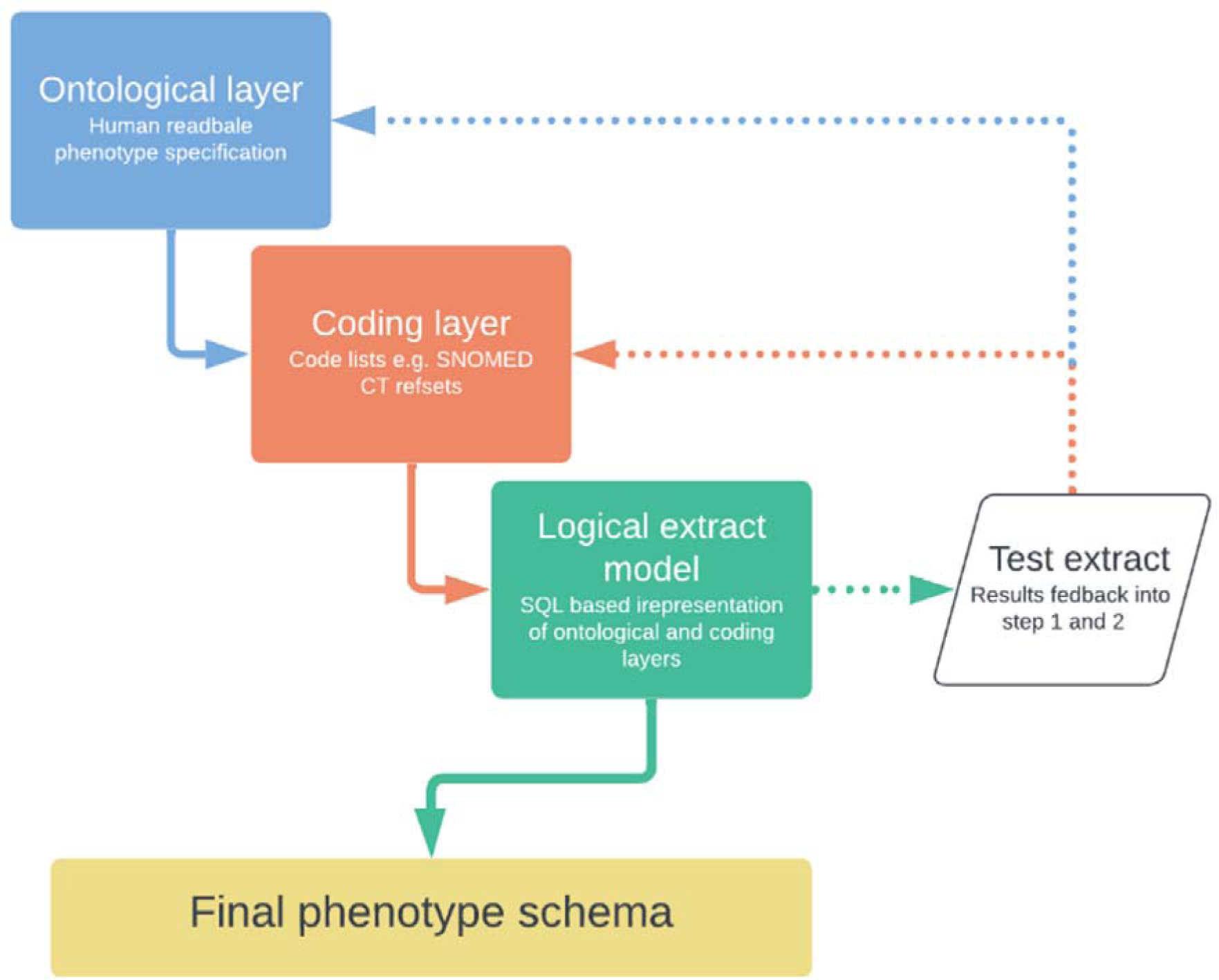
Existing three-layered approach for defining phenotypes at the RSC. **Ontological layer:** human-readable phenotype specification, may utilise diagnostic criteria, symptom examination findings, test result and therapies. **Coding layer:** Creation of code lists typically using SNOMED CT. **Logical extract model:** Structured Query Language (SQL) scripts are manually developed based on ontological and coding layer. **Test extract:** Assessed for veracity modifications are fed back to ontological and coding layers. The phenotype schema can then be finalised.

We have previously defined important phenotypes such as pregnancy, chronic kidney disease, social prescribing interventions, and long COVID with Web Ontology Language (OWL) using the Protégé software.[10,11,12] These phenotypes were then uploaded to the public phenotype libraries Bioportal and PhenoFlow.[13]

### Case for changing our approach to developing phenotypes

We are changing our approach to developing phenotypes as, firstly, we found OWL excellent for semantic precision, but sometimes challenging to align with clinical and project reasoning. Secondly, in the coding layer we had previously defined refsets ‘extensionally’. This is where each code is explicitly listed as a member of the sets. We now plan to make better use of the SNOMED expression constraint language (ECL).[14,15,16] SNOMED CT is designed to be machine processable and its ECL allows a rule-based or ‘intensional’ approach to developing refsets that utilise SNOMED’s polyhierarchical subtypes and supertypes.[17,18] The logic used to define intensional refsets is more explicit.

### Evaluation of Phenotype Execution and Modelling Architecture (PhEMA)

Phenotype Execution and Modelling Architecture (PhEMA) is a standards-based, modular architecture for developing phenotype algorithms, including their validation, execution and dissemination developed for real world studies using CMR data.[19] PhEMA uses aspects of Health Level 7’s Fast Healthcare Interoperability Resources (FHIR) framework including the use of Clinical Quality Language (CQL). CQL is a structured way of expressing phenotype logic and code lists and allows expression and sharing of phenotype logic, it is designed to be human and machine readable. [20,21].

This study aimed to evaluate PhEMA’s CQL-based logic model and creation of Health Level 7 (HL7) Fast Healthcare Interoperability Resources (FHIR) valuesets, into the RSC’s three-layered approach to phenotyping, replacing OWL or unstructured phenotype definitions. We adapted our approach to incorporate PhEMA elements and then aimed to develop phenotypes for three purposively selected use cases: type 2 diabetes mellitus (T2DM), incident influenza-like illness (ILI), and excessive alcohol consumption.

## Method

### The three phenotype use cases

We selected three dissimilar use cases that we anticipated would require different logic models. The three we selected were: (1) Type 2 diabetes mellitus (T2DM), (2) Excessive alcohol use, and (3) Incident cases of influenza-like illness (ILI). We compared the new phenotypes with those produced using our current process, and where a variable is new looked for external validation.

#### Use case 1: Type 2 diabetes mellitus

We chose the T2DM phenotype due to our extensive experience with CMR-based T2DM studies and its frequent use as a covariate in other research.[22,23,24] Our involvement extends to examining misdiagnosis, miscoding, and misclassification within CMRs.[25,26,27] Phenotyping T2DM is challenging because the diagnosis can be derived through numerous clinically coded events, including diagnostic codes, laboratory results or the prescribing of medication. Additionally, T2DM may resolve through lifestyle interventions and bariatric surgery, but there is potential later relapse.[28,29,30] The RSC has a well-established but complex phenotyping algorithm based on our existing three-layered approach.[31] To evaluate PhEMA’s potential integration, however, we will use a simplified version of our current T2DM phenotype.

#### Use case 2: Excessive alcohol use

The alcohol use case phenotype is an exemplar of a sociodemographic risk factor. We identified a variety of definitions of excessive alcohol use and chose one based on consumption levels rather than questionnaires about the effects of drinking. The NHS defines excessive consumption of alcohol as more than 14 units of alcohol per week.[32]

#### Use case 3: Incident influenza like illness

The World Health Organization (WHO) recommends surveillance of the respiratory clinical syndrome ILI as a marker of community spread of respiratory infections. Tracking the incidence of ILI is important because in respiratory surveillance systems around half of ILI clinical cases have laboratory confirmed influenza.[33] Alongside other surveillance indicators ILI surveillance should be part of the mosaic of measures used to flag an emergent pandemic.[34,35] The RSC provides weekly ILI incidence figures to UKHSA in its weekly report.

### Modifying our three-step ontological process for defining a phenotype in real-world data

Our modified three-step approach to embrace PhEMA principles is shown in Figure 2. The logic model and coding layer use the PhEMA processes with logic model expressed in CQL. The PhEMA logic model includes operational factors in addition to clinical concepts. Operational factors describe non-clinical factors that influence CMR coding and may relate to the data entry environment or systems of practice. For instance, variations in the use of specific codes might be influenced by contractual requirements and incentives; this context is essential for our approach. Additionally, we alter the criteria to adjust the sensitivity or specificity of the extraction according to the aims of the study.[36] The logical data extraction step is external to the PhEMA process. Here we generate the data output and check the internal and external validity of new phenotypes.

**Figure 2:**
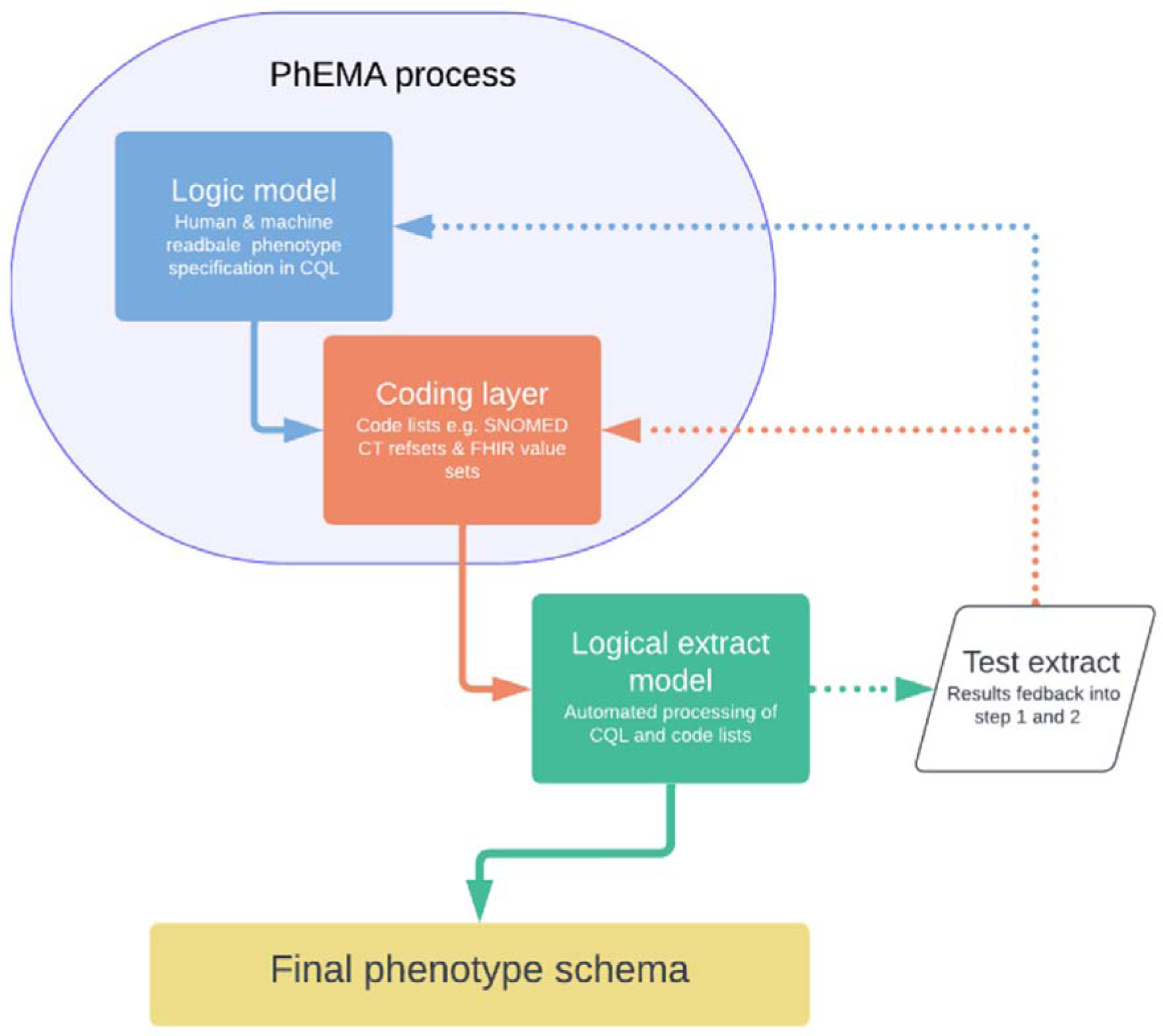
Revised three-step model for developing phenotypes at the RSC. **Logic model:** Define clinical and operational logic using Clinical Quality Language (CQL). **Coding layer:** Create intensional refsets using in-house SNOMED CT helper tool (Supplementary Figure S1). SNOMED refsets are converted to Fast Healthcare Interoperability Resources (FHIR) valuesets for CQL compatibility. **Logical extraction model:** Automated processing of machine readable PhEMA output to extract data from CMR. **Test extract:** Assessed for veracity modifications are fed back to PhEMA layers. Internal and external validation processes.

### Step 1: Describing the logic model using Clinical Query Language (CQL)

We used the PhEMA work bench to define the phenotype logic model for each of our three use cases in CQL.[37] Subsequently we used Visual Studio Code from Microsoft, with the CQL extension which provides syntax highlighting and debugging tools.[38,39] Our CQL code is based resources developed in the FHIR and PhEMA helper libraries.[40,41] The resources produced under the PhEMA and Clinical Quality Framework (CQF), including CQL, contain a set of examples that can be used to generate the majority of commonly used phenotypes. We found the HL7 Confluence community workspace, the most accessible route to get started.[42]

### Step 2: Defining the FHIR valuesets and SNOMED CT refsets

SNOMED refsets were initially created using our SNOMED Helper Tool (Supplementary Figure S1). SNOMED’s ECL allows sets of concepts to be defined intensionally from their relationships.[43] Most often this is a hierarchical relationship where a high-level concept (known as a supertype) is explicitly defined and all lower level concepts (known as subtypes), whose meanings are subsumed by the higher concept, are included by inference.14,15,16] Conversely, concepts may be excluded by specifying a minus supertype and none of its subtypes will be included. The SNOMED ECL functions we used included “Child or Self of” (written childOrSelfOf) where a clinical term and its child codes are included.[44] SNOMED refsets were then converted to FHIR valuesets.

FHIR valuesets are a set of clinical terms and associated metadata that are compatible with multiple code systems, including intensional SNOMED CT expressions or refsets. Metadata can include the coding or classification system used, the valueset provenance and lifecycle management, such as marking a valueset as experimental.[45] A FHIR valueset can be marked as a draft, or at end of life, be formally retired whilst preserving the content.[46]

### Step 3: Describing the logical extract model

We used the developed phenotypes to extract the associated cohort of patients from the RSC SQL-based CMR. We did this by manually converting CQL phenotype logic to SQL. We plan to automate this step in the future to take full advantage of the machine-readable nature of CQL. From the extracted data, we then calculated summary statistics for each of the use cases.

#### T2DM

We compared the prevalence of T2DM in the current adult (>18 years) RSC population to that of a recent cohort that used our previous phenotype. Additionally, we compared the prevalence in all age groups to that of the estimated UK diabetes prevalence using data from Diabetes UK and the Office for National Statistics (ONS).

#### Excessive alcohol use

We assessed excessive alcohol consumption prevalence using qualitative SNOMED codes, then combined them with quantitative SNOMED codes, specifically the units of alcohol consumed, and determined relative percentage increase in prevalence with the addition of quantitative codes.

#### Influenza like illness

We compared the incidence of ILI for ISO weeks 26 to 38 of 2023, the most recent data available, using both the old and new phenotypes. We used these dates as a convenience sample. We report the median, lower and upper quartiles of the weekly incidence of ILI by age band for this period. The age bands used are consistent with those presented in the weekly surveillance report to UKHSA.

### Ethical considerations

The variables curated in this work are all required for our sentinel surveillance, approved under Regulation 3 of the Health Service (Control of Patient Information) Regulations 2002, and reviewed and approved annually by the UKHSA Caldicott Guardian.[47]

## Results

### Use case 1: Type 2 diabetes mellitus

#### Logic model

Clinical terms that inferred a diagnosis of T2DM were identified intensionally from the SNOMED hierarchy. For this simplified diabetes phenotype, we did not use blood glucose levels or prescription of medication to indicate a diagnosis of T2DM. We also identified individuals whose diabetes had resolved and excluded them from the final cohort. For operational logic we included process of care codes which imply a diagnosis of T2DM, for example, “Type 2 DM review”, (STCID: 279321000000104). The CQL code can be seen in Figure 3.

**Figure 3.**
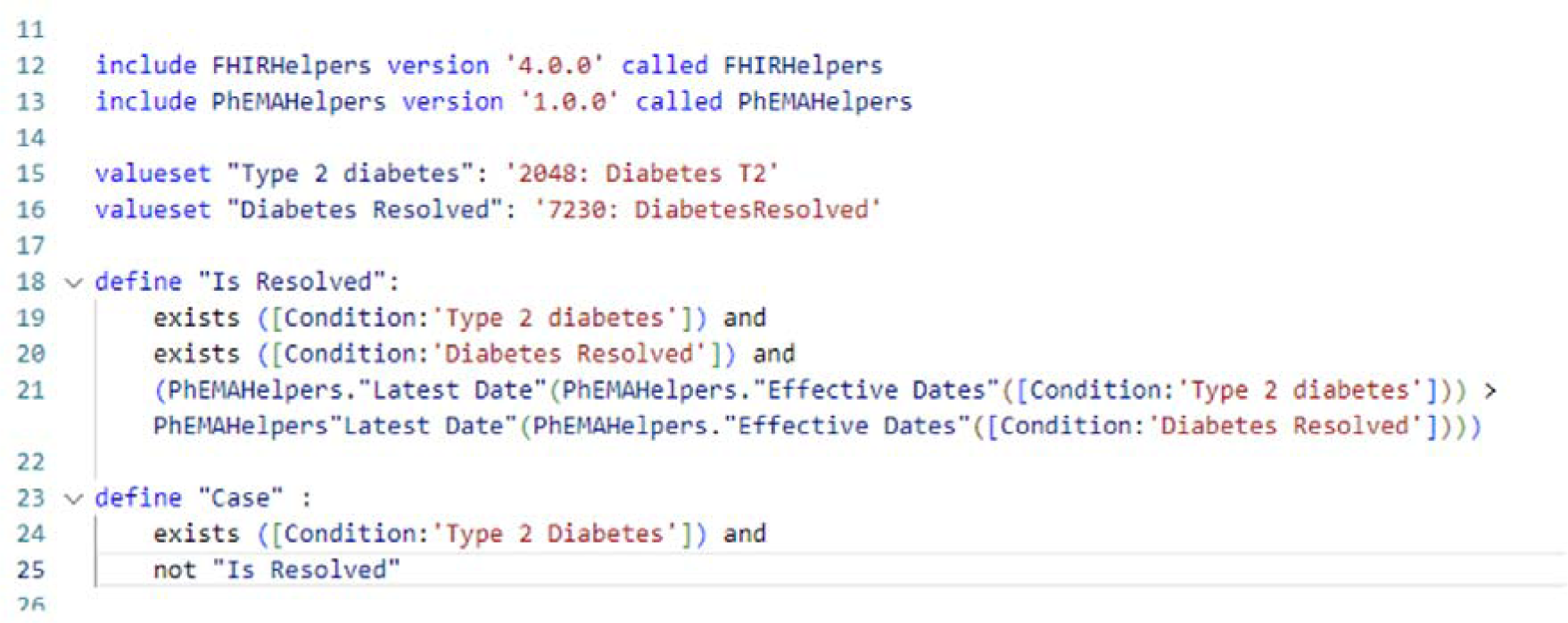
Clinical quality language (CQL) logic model for our type 2 diabetes (T2DM) phenotype. Individual is coded as resolved if resolved date is greater than diagnosis date.

#### Coding layer

We created two SNOMED CT refsets, the first consisting of codes implying the diagnosis of diabetes and the second implying that the diagnosis had resolved. The first refset included the diagnostic codes such as “Diabetes mellitus type 2” (SCTID: 44054006) and all of its descendant codes. We additionally used single relevant clinical terms such as “Type 2 diabetic on insulin” (SCTID: 24471000000103) and other concepts and their relevant child concepts. Subtypes make up the bulk of the definition - 97% of the concepts in the final, expanded, extensional set arose from the “childOrSelfOf” definitions (Supplementary Textbox S1).

#### Logical extract model

In individuals over 18 years of age, the RSC CMR’s diabetes prevalence using the new phenotype was 7.3%. This is slightly higher than the 7.2% prevalence from a 2021 cohort study on sodium-glucose co-transporter-2 inhibitor prescriptions in primary care, which utilized the old RSC phenotype and 2019 RSC data.[48] For a national comparison, we incorporated data for those under 18. The prevalence across all age groups with the new phenotype was 5.9%, in contrast to an approximate 5.3% derived from Diabetes UK and the Office for National Statistics data.[49]

### Use case 2: Excessive alcohol use

#### Logic model

Using only clinical logic we identified individuals with evidence of consumption of greater than 14 units of alcohol per week. Firstly, we looked for qualitative codes with a rubric that includes the specific number of units consumed, for example “moderate drinker 3-6u/day”, (SCTID: 160576006). We then identified codes allowing input of qualifying values, in this case the specific number of units consumed by an individual. Our final logical model for excessive alcohol use (Figure 4), with the CQL logic in a supplementary file.

**Figure 4.**
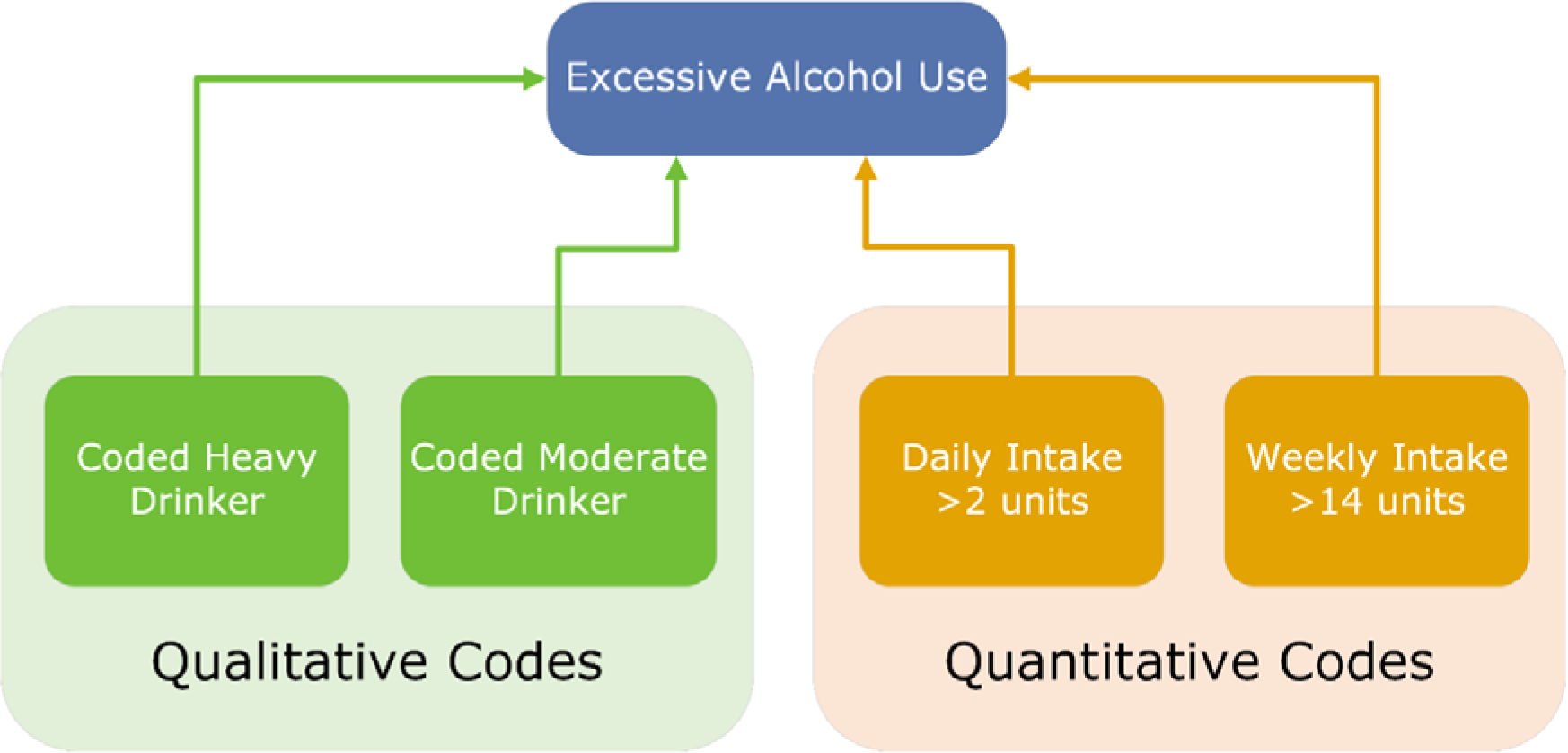
Clinical concepts used to define the excessive alcohol use valueset. Including qualitative codes and quantitative codes that represent the number of units of alcohol consumed in the specified time period.

#### Coding layer

We created four valuesets which combined to produce our phenotype, two containing qualitative codes and two contained codes with associated qualifying values (quantitative codes). The qualitative value sets contain concepts from the “findings” section of the SNOMED CT hierarchy. With one valueset defining heavy drinkers including “Heavy drinker 7-9u/day” and “Very heavy drinker >9u/day” codes. The second valueset contains the single concept for moderate drinker. Two valuesets will contain “observable entity” concepts from the UK SNOMED CT Extension: “Alcohol units consumed per day” and “Alcohol units consumed per week”. The CQL for excessive alcohol consumption can be seen in Supplementary Textbox S2.

#### Logical extract model

The overall prevalence of excessive alcohol consumption in the RSC CMR using only qualitative SNOMED codes was 1.2%. The addition of quantitative codes to the phenotype increased the prevalence to 1.9%, a 58.3% relative increase although this is still considerably below the Health Survey data for England which reported 21% of adults drinking more than 14 units weekly.[50] Table 1 shows the breakdown of prevalence by age band. The increase in prevalence at age forty is likely due to the NHS Health Check which includes questions about alcohol use and is offered to all registered patients every five years from age 40 to age 75.[51]

**Table 1:**
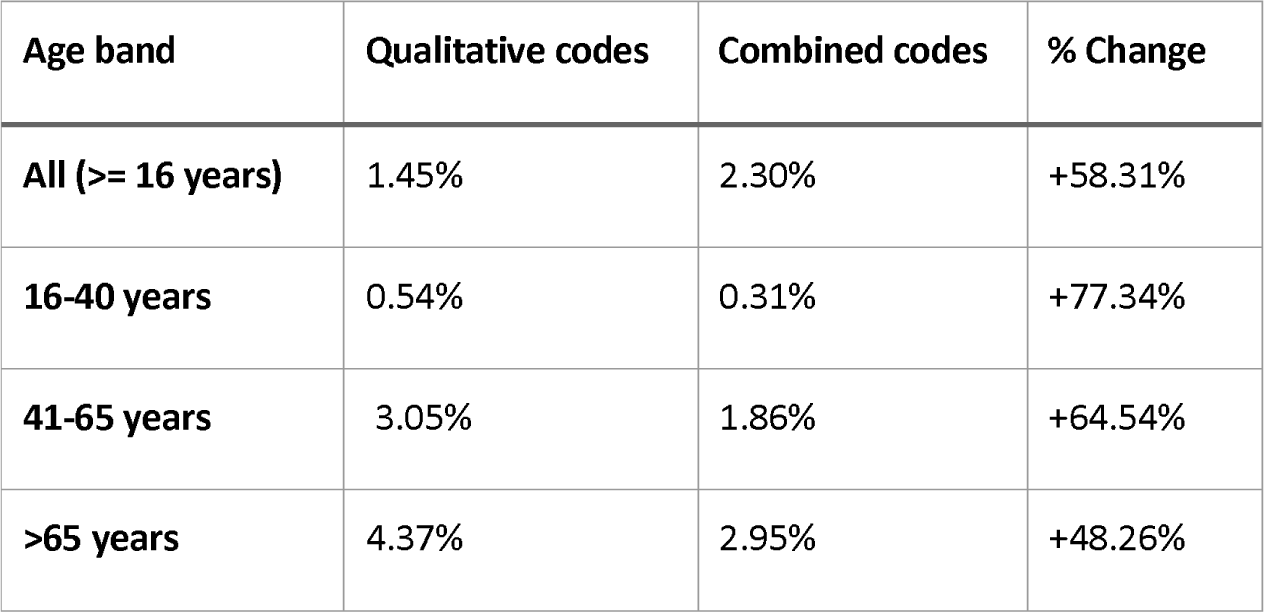
Prevalence of excessive alcohol consumption in the RSC in those 16 years or older. **a**: Prevalence using only qualitative codes. **b**: Prevalence using qualitative and quantitative codes. **d**: the percentage increase in prevalence through use of combined codes.

| Age band | Qualitative codes | Combined codes | % Change |
| --- | --- | --- | --- |
| All (>= 16 years) | 1.45% | 2.30% | +58.31% |
| 16-40 years | 0.54% | 0.31% | +77.34% |
| 41-65 years | 3.05% | 1.86% | +64.54% |
| >65 years | 4.37% | 2.95% | +48.26% |

### Use case 3 Influenza-like Illness

#### Logic Model

As ILI is an acute clinical syndrome that may recur multiple times in the same individual, we aimed to identify weekly incident cases. This contrasts with the previous use cases in which we were identifying the prevalence of a condition. We developed an approach to distinguish between new and repeat presentations of the same ILI episode using the PhEMA framework. We identified ILI clinical terms recorded more than 28 days, from the previous recorded event (Supplementary file ILI.cql). As this requires use of date values within CQL we used the PhEMAHelpers library which contains functions to simplify representation of dates.

#### Coding layer

We created a single valueset from the SNOMED CT identifying conceptually different areas within SNOMED CT which met our clinical criteria for ILI. These included the following codes and their subtypes: “Influenza-like illness”, “Influenza-like symptoms” and “Influenza”. Additionally, we excluded high-level concepts within the hierarchy such as “Healthcare associated infections” which excludes “Healthcare associated influenza disease” from within the “influenza” parent code.

#### Logical extract model

Between ISO weeks 26 and 38 of 2023, the median weekly incidence of ILI using the new phenotype definition was 1.11 cases per 100,000 people. In contrast, using the old phenotype definition, it was 0.95 cases per 100,000 people. Table 2 shows the weekly incidence of ILI by age band.

**Table 2:** Median weekly incidence of ILI in the RSC CMR for ISO weeks 26 to 38 of 2023 using both the new and old phenotypes. Values denote medians, with lower and upper quartiles enclosed in brackets.

| Age band | New phenotype | Old phenotype |
| --- | --- | --- |
| All ages | 1.11 (0.66-1.44) | 0.95 (0.60-1.25) |
| <15 years | 0.54 (0.34-0.68) | 0.53 (0.25-0.69) |
| 15-64 years | 1.51 (1.31-1.65) | 1.41 (1.16-1.54) |
| >65 years | 1.16 (1.00-1.22) | 1.02 (0.68-1.16) |

## Discussion

### Principal Findings

We have demonstrated that we can effectively use PhEMA to curate phenotypes - creating valuesets and more shareable logic. SNOMED refsets created intensionally take advantage of SNOMED’s consistent hierarchy and ECL. Using CQL to express clinical and operational logic was comprehensible to clinicians and epidemiologists in our team and will be adopted. We have demonstrated this across three different use cases. Across all these use cases the case acquisition increased. We anticipated that the prevalence of excessive alcohol would increase, as we were adding qualitative clinical terms. However, it was not anticipated for T2DM where we added the capability to go into remission or in ILI where we added a temporal constraint to help ensure that we only counted incident cases. Our interpretation is that the intension use of SNOMED’s ECL accounted for this increase.

### Implications of the finding

We will migrate our approach to curating variables from OWL, where we were more challenged to express clinical logic, to PhEMA and CQL. The separation of clinical logic from administrative, largely health system constraints, was considered helpful. Standardising our valuesets using the FHIR metadata improves our ability to share and version control our curated variables. This is a step towards phenotypes that are readily machine processable. Intension use of SNOMED’s ECL may be important in ensuring that all relevant clinical terms within that hierarchy are captured.

### Comparison with the literature

There have been several attempts to allow sharing of phenotypes.[52,53,54] These have used their own formats for expressing the criteria for extracting data from the medical record. Logic is typically described in free text or expressed using general scripting languages with an implementation which is closely tied to the underlying data structure and storage used by the researcher.

In the UK the leading organisations promoting shared phenotypes and code lists have been Health Data Research UK and OpenSafely respectively.[55,56] However, neither of these include interoperable expressions of clinical logic.

The US National Library of Medicine distributes valuesets through the Valueset Authority Center,[57] but the NHS England Terminology Server lacks the former’s scope.[58] Health records and SNOMED CT differ significantly between in the USA and the NHS.

We unambiguously accepted the rigor of the SNOMED hierarchy, and whilst vastly better than the inconsistencies of the UK’s previous Read codes, there are limitations and questions as to whether they are sufficiently quality assured.[59,60]

### Strengths and imitations

The research group have had long experience of curating variables across the range of clinical terminologies used in the English NHS. We have shown that phenotypes can be expressed and distributed in a way that is comprehensible to both humans and computers and that implementing the PhEMA Framework is technically and practically feasible.

Whilst we have met three of the desiderata that Chapman[4] expressed for the creation of a next generation phenotype library, the many gaps that remain are a limitation of this process. We demonstrated this process using the hierarchies of SNOMED CT, but it would apply to less ontologically complex terminologies such as ICD or OPCS. Our conclusions about the interpretability of CQL are based on the opinions of the authors and no formal consensus building or qualitative assessment was conducted.

We will need considerable effort to convert our large library of curated variables into PhEMA format. The lack of an open valueset authority in the UK, as exists in USA, limits our ability to collaborate and share valuesets.

## Conclusion

We have shown that we can express phenotypes using the principles of the PhEMA framework for the identification of patients with chronic disease, the assessment of observations and for surveillance of communicable disease using international health data standards.

It was feasible and practical to distribute FHIR valuesets using intensional definitions which can be compiled to extensional lists by terminology servers. This approach also increased case finding.

We have described the logic unambiguously using CQL, a FHIR specification, in a way which is understandable for humans as well as potentially computable.

## Supporting information

Supplementary Textboxes

CQL files

## Data Availability

All data produced in the present work are contained in the manuscript and additional files

## Acknowledgements

Patients who allow data sharing registered with practices who are member of the Royal College of General Practitioners (RCGP) Research and Surveillance Centre (RSC). EMIS Magentus, TPP, and InPractice Systems for facilitating pseudonymised data sharing.

## CONRTIBUTIONS

SdeL and GJ drafted the text and GJ assisted by DK, RW and AF created the valuesets. WE, RB, WH performed the data extraction and analysis. RB and Dr John Williams created the SNOMED CT Helper Tool. All the authors read and reviewed the manuscript.

