## Supplementary Figure S1 for "Phenotype Execution and Modelling Architecture (PhEMA) to support disease surveillance and real-world evidence studies: English sentinel network evaluation"

### **Supplementary file**

**Figure S1:** Helper Tool used to facilitate the development of SNOMED CT refsets

The upper figure displays the type two diabetes (T2DM) refset. The lower annotated figure demonstrates how SNOMED CT supertypes and subtypes are included or excluded using the Helper Tool.

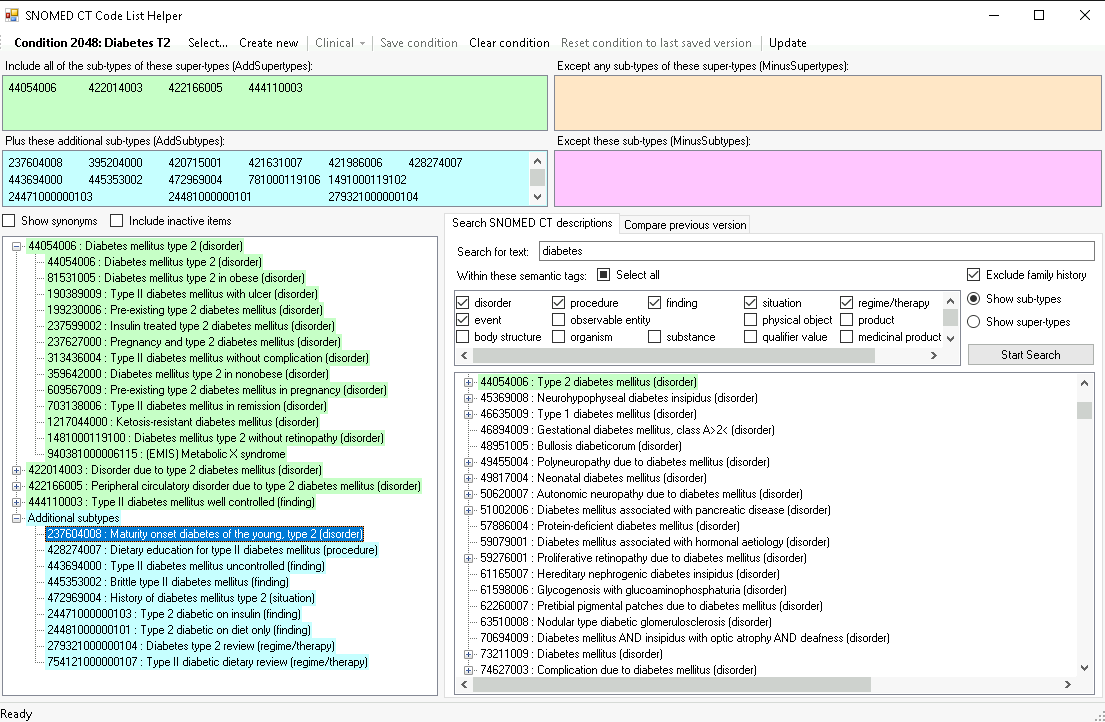

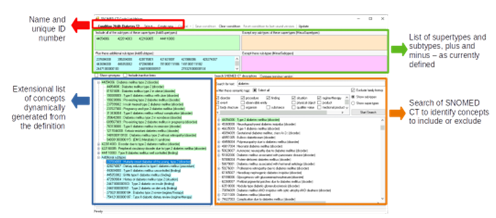
