## Supplementary Textboxes for "Phenotype Execution and Modelling Architecture (PhEMA) to support disease surveillance and real-world evidence studies: English sentinel network evaluation"

**Textbox S1:** The T2DM valueset

Valueset for type 2 diabetes use case, using SNOMED CT expression constraint language. This is the long form of the ECL which is easier for humans to read. The definition contains 13 rules which, when run against the current UK edition of SNOMED CT produced a list of 195 concepts including “History Supplement” to include inactivated SNOMED CT concepts which may still be present in the CMR and are considered equivalent to selected active conceptslxi.

- childOrSelfOf 44054006 |Diabetes mellitus type 2| {{ + HISTORY ( 900000000000527005 |SAME AS association reference set| ) }} *
- childOrSelfOf 422014003 |Disorder due to type 2 diabetes mellitus|
- childOrSelfOf 422166005 |Peripheral circulatory disorder due to type 2 diabetes mellitus|
- childOrSelfOf 444110003 |Type II diabetes mellitus well controlled|
- 237604008 |Maturity onset diabetes of the young, type 2|
- 428274007 |Dietary education for type II diabetes mellitus|
- 443694000 |Type II diabetes mellitus uncontrolled|
- 445353002 |Brittle type II diabetes mellitus|
- 472969004 |History of diabetes mellitus type 2|
- 24471000000103 |Type 2 diabetic on insulin|
- 24481000000101 |Type 2 diabetic on diet only|
- 279321000000104 |Diabetes type 2 review|
- 754121000000107 |Type II diabetic dietary review|
- *History used in all lines omitted for clarity

**Textbox S2:** The four valuesets used in the excessive alcohol use phenotype. Heavy drinker uses supertypes but the extensional set is only four concepts in total. The other three valuesets contrain a single concept each.

Heavy drinker

childOrSelfOf 86933000 |Heavy drinker|

childOrSelfOf 228279004 |Very heavy drinker|

Moderate drinker

160576006 |Moderate drinker – 3-6u/day|

Alcohol Units Consumed Per Day

1082631000000102 |Alcohol units consumed per day|

Alcohol Units Consumed Per Week

1082641000000106 |Alcohol units consumed per week|

**Textbox S3:** The influenza-like illness valueset

- childOrSelfOf 6142004 |Influenza|
- childOrSelfOf 78046005 |Myocarditis caused by influenza virus|
- childOrSelfOf 195929004 |Influenza with gastrointestinal tract involvement |
- childOrSelfOf 309789002 |Encephalitis caused by influenza|
- 95891005 |Influenza-like illness|
- 315642004 |Influenza-like symptoms|
